# Neonatal Resuscitation Training in Australian Public Birthing and Newborn Care Facilities: A National Cross-Sectional Survey

**DOI:** 10.64898/2026.10.21.26366083

**Authors:** Stephanie Morakeas, Mark Tracy, Nicholas Williams, Marta Thio

## Abstract

**Background:** The stabilisation of infants at birth is vital for survival and relies on clinicians being appropriately trained in neonatal resuscitation. The effectiveness of resuscitation is influenced by the skills, knowledge and equipment available to the responding clinician. In Australia, the absence of a standardised neonatal resuscitation training programme may contribute to variations in training practices. This study aimed to describe current training practices across Australian public services and explore educators’ perceptions of opportunities for national standardisation.

**Methods:** A cross-sectional survey was distributed to educators and representatives from birth and newborn care public facilities across Australia. A questionnaire was developed to gain knowledge on how newborn resuscitation is taught detailing staffing, equipment and training requirements.

**Results:** 197 completed responses were received. Responses were received from all Australian states and territories and levels of maternity/neonatal services. Accreditation in neonatal resuscitation was required by 173 (87.8%) of respondents’ programmes with training occurring at annual (n=95, 47.7%) or more frequent (n=21, 10.7%) intervals. NeoResus (n=134, 67.5%), Neonatal Advanced Life Support (n=45, 22.8%) and Newborn Life Support Training (n=41, 21.3%) were the most popular training programmes. Substantial variation was identified in equipment used for training. Most respondents (n=174, 88.3%) reported a need for a standardised national training programme.

**Conclusions:** Neonatal resuscitation training is widely available in Australian public facilities, but reported requirements, programmes, content, assessment methods and training resources vary. These findings support the development of standardised neonatal resuscitation training to promote consistent preparedness across Australian healthcare settings.

- **What is already known on this topic** Regular neonatal resuscitation training is recommended by international guidelines and is associated with improved neonatal outcomes. Previous Australian studies have identified a variation in training access and requirements, but contemporary national data are lacking.
- **What this study adds – summarise what we now know as a result of this study that we did not know before** This study provides the first national baseline of neonatal resuscitation training practices across Australian healthcare services. It identifies substantial differences in training requirements, programme delivery and educational resources, providing support and information to inform a standardised national approach.
- **How this study might affect research, practice or policy – *summarise the implications of this study*** The findings provide evidence to support development of coordinated national approach to neonatal resuscitation training and may inform future standards, educational policy and research to improve education.

## Introduction

Newborn resuscitation is an uncommon event for individual clinicians, creating a significant challenge for maintaining clinical proficiency. In Australia, 21% of births require some form of active resuscitation.^1^ While interventions such as suction (7.0%), oxygen administration (3.8%), continuous positive airway pressure (10.6%) and intermittent positive pressure ventilation (4.4%) are encountered more frequently, advanced procedures such as intubation (0.9%) and external cardiac compressions (ECC) (0.2%) are performed infrequently.^1^

Resuscitation outcomes vary according to place of birth with premature infants born in non-tertiary centres experiencing higher mortality than those born in tertiary centres.^2 3^ Whilst efforts are made to facilitate delivery of high-risk infants in tertiary hospitals, this is not always possible. For clinicians working in centres where exposure to neonatal resuscitation may be limited, structured education and training play an important role in maintaining the knowledge, skills and teamwork required for effective resuscitation. Evidence suggests that neonatal resuscitation training improves clinical outcomes, with a meta-analysis demonstrating a 42% reduction in first-day neonatal mortality following training programmes,^4^ findings that are supported by a Cochrane review reporting reduced mortality at both 24 hours and 7 days of life.^5^

International guidelines emphasise the importance of ongoing training. The International Liaison Committee on Resuscitation (ILCOR) recommends that training should be recurrent and undertaken more frequently than annually.^6^ Similarly, the Australian and New Zealand Committee on Resuscitation (ANZCOR) provides clinical guidance supporting recurrent training and highlights the need for organised programmes to develop and maintain resuscitation knowledge, skills and teamwork.^7^ In Australia, whilst the Australian Resuscitation Council (ARC) provides clinical guidance, it does not oversee implementation of neonatal resuscitation training. As a result, training frequency, programme structure, accreditation requirements, and access to training are not standardised nationally. Training recommendations vary between jurisdictions, with state health departments recommending refresher training every one to two years.^8^ The training programmes recommended or mandated also differ between states and professional organisations.^9 10^

Previous Australian studies have identified inequities in access to neonatal resuscitation training. Foster et al reported lower training exposure and confidence among clinicians working in rural and non-tertiary centres compared with tertiary hospitals in NSW and the ACT.^11^ Similarly, a review of neonatal resuscitation training requirements for Australian midwives found substantial variation in state-based polices, training programmes, teaching methods and training frequency, with only four states mandating neonatal resuscitation training through statewide guidelines.^9^ These studies were limited to specific jurisdictions, professional groups or policy reviews and do not provide contemporary national data describing the delivery of neonatal resuscitation training across Australian healthcare services.

This study aims to provide a contemporary national overview of neonatal resuscitation training across Australian public hospitals through a national survey of neonatal resuscitation educators. Specifically, the study sought to characterise programme frequency, content, teaching methods and the resources available to support training delivery and explore educators’ perceptions of opportunities for national standardisation.

## Methods

### Study Design

A cross-sectional survey was conducted to establish a national baseline for newborn resuscitation training practices across Australian public hospitals. Survey questions were developed, reviewed and pretested by an expert multidisciplinary panel consisting of neonatologists, nurse practitioners and nurse educators. A 30-item questionnaire was developed in REDCap (Research Electronic Data Capture, Office for Health and Medical Research) and consisted of three sections containing multiple-choice, multi-select and open-ended questions (questionnaire is available in Supplementary Appendix 1). The questionnaire examined the content and implementation of neonatal resuscitation training programmes, the equipment and educational resources utilized, alignment with current ANZCOR resuscitation training guidelines and the approaches used to evaluate programme effectiveness. Survey reporting followed the CROSS checklist.^12^

### Participants

Participants were healthcare professionals involved in neonatal resuscitation training at Australian public hospitals, including medical, nursing and midwifery staff. Public hospitals providing maternity and/or neonatal services across all levels of care were included. In the absence of national service level classification system, facilities were categorised according to the NSW maternity service capability framework.^13^ A sampling frame of eligible healthcare professionals in public healthcare facilities across metropolitan, regional and rural Australia was established through the compilation of an email distribution list. Where possible a single representative from each hospital was identified, typically an educator responsible for newborn resuscitation training. In some jurisdictions, district level educators overseeing training at multiple hospitals were invited to participate and either complete the survey themselves or distribute it to appropriate personnel. As multiple contacts were identified for some hospitals it is possible that more than one survey response was submitted from the same site.

### Data collection

The survey was distributed to the email list electronically using REDCap. Participants were provided with a four-week period to complete the survey and received reminder emails at two weeks and shortly before survey closure, resulting in a total of three survey invitations. Respondents were asked to provide responses based on practices at their current workplace. No incentives were provided for participation.

Survey responses were collected anonymously using REDCap. As the survey was distributed via an open survey link to facilitate participation of relevant individuals, it was not possible to prevent multiple submissions.

### Outcomes

The primary outcome was the characterisation of neonatal resuscitation training practices in Australian public hospitals. The secondary outcomes included training program type and implementation, equipment, educational resources, evaluation of strategies used to assess program effectiveness and alignment with ANZCOR guidelines.

### Sample Size

Based on estimates of Australian public maternity and newborn care facilities, approximately 254 hospitals were considered eligible for participation. A response rate of 60% of facilities (approximately 153 hospitals) was anticipated based on a previous Australian survey.^14^ Educators representing 232 facilities were identified and survey invitations were distributed via 222 email contacts. Multiple individuals were contacted for some facilities, while some educators were responsible for training across multiple facilities. As survey recipients were encouraged to forward the survey link to relevant colleagues and responses were anonymous, the total number of individuals receiving the invitation was unknown. Consequently, a participant or facility level response rate could not be calculated.

### Statistical Analysis

Data were exported from REDCap into StataMP (version18) for descriptive statistical analysis. Categorical variables were summarised using frequencies and percentages. Free-text responses were analysed using inductive content analysis to identify common themes related to training practices and perceived needs for national standardisation. To present data in a simplified format some data were grouped as tertiary (maternity/neonatal service > Level 4) and non-tertiary units (maternity/neonatal service ≤ Level 4).^13^ Partially completed responses were if sufficient data were provided to address the study objectives, however responses were excluded from analysis if the respondent did not select an Australian state or territory.

### Ethics Approval

Ethics approval was obtained from the Western Sydney Local Health District Human Research Ethics Committee (2025/ETH00068) prior to survey dissemination. Participation was voluntary with responses collected anonymously. Completion of the survey implied informed consent.

## Results

### General Characteristics

A total of 199 survey responses were received between September and October 2025. Two incomplete responses were excluded, leaving 197 completed responses included in the final analysis (99% completion proportion). Participant demographics are summarised in Table 1.

**Table 1:** Participant demographics.

| **Participants (n=197)** | **Number of Respondents (%)** |
| --- | --- |
| **Role** |  |
| Senior Medical Staff | 22 (11.2%) |
| Nursing Staff | 16 (8.1%) |
| Midwifery Staff | 86 (43.7%) |
| Nursing/Midwifery Educators | 73 (37.0%) |
| **Clinical Experience in Teaching Newborn Resuscitation** |  |
| 0 years | 15 (7.6%) |
| <1 year | 17 (9.3%) |
| 2-5 years | 66 (36.6%) |
| 5-10 years | 42 (23.1%) |
| >10 years | 57 (31.3%) |
| **Location** |  |
| QLD | 56 (28.4%) |
| NSW | 54 (27.4%) |
| VIC | 41 (20.8%) |
| WA | 19 (9.6%) |
| SA | 15 (7.6%) |
| TAS | 5 (2.5%) |
| NT | 4 (2.0%) |
| ACT | 3 (1.5%) |
| **Maternity/Neonatal Service Level\*** |  |
| Level 1 (No planned maternity/neonatal service) | 6 (3.0%) |
| Level 2 (newborns ≥37 weeks) | 32 (16.2%) |
| Level 3 (newborns ≥34 weeks) | 28 (14.2%) |
| Level 4 (newborns ≥32 weeks) | 49 (24.9%) |
| Level 5 (intensive care for critically ill newborns including extreme preterm infants) | 39 (19.8%) |
| Level 6 (neonatal surgical and medical subspecialty services) | 43 (21.8%) |
| * NSW Health Guide to Role Delineation of Clinical Services (Supplementary Appendix 2)<br>Note: Distribution of participants by state/territory and maternity/neonatal service level is presented in Supplementary Appendix 1 Table 1. |  |

### Training Requirements and Delivery

Accreditation in neonatal resuscitation was specified as required by 173 (87.8%) of respondents. Respondents’ institutions training intervals are presented in Figure 1, two responses indicating ‘Other’ reported that their institution had no set training interval or did not require regular training only a competency assessment. Training duration is presented in Figure 2. Pre-course work was required by 91.9% of respondents’ programs. Training is performed onsite (n=190, 96.4%), offsite (n=26, 13.2%), online (n=54, 27.4%), unsure (n=1, 0.5%). Training was facilitated by doctors/nurse practitioners (n=10, 5.1%), nurses/midwives (n=64, 32.5%), both (n=100, 50.8%), other (n=23, 11.7%). Appendix Table 1.

**Figure 1:**
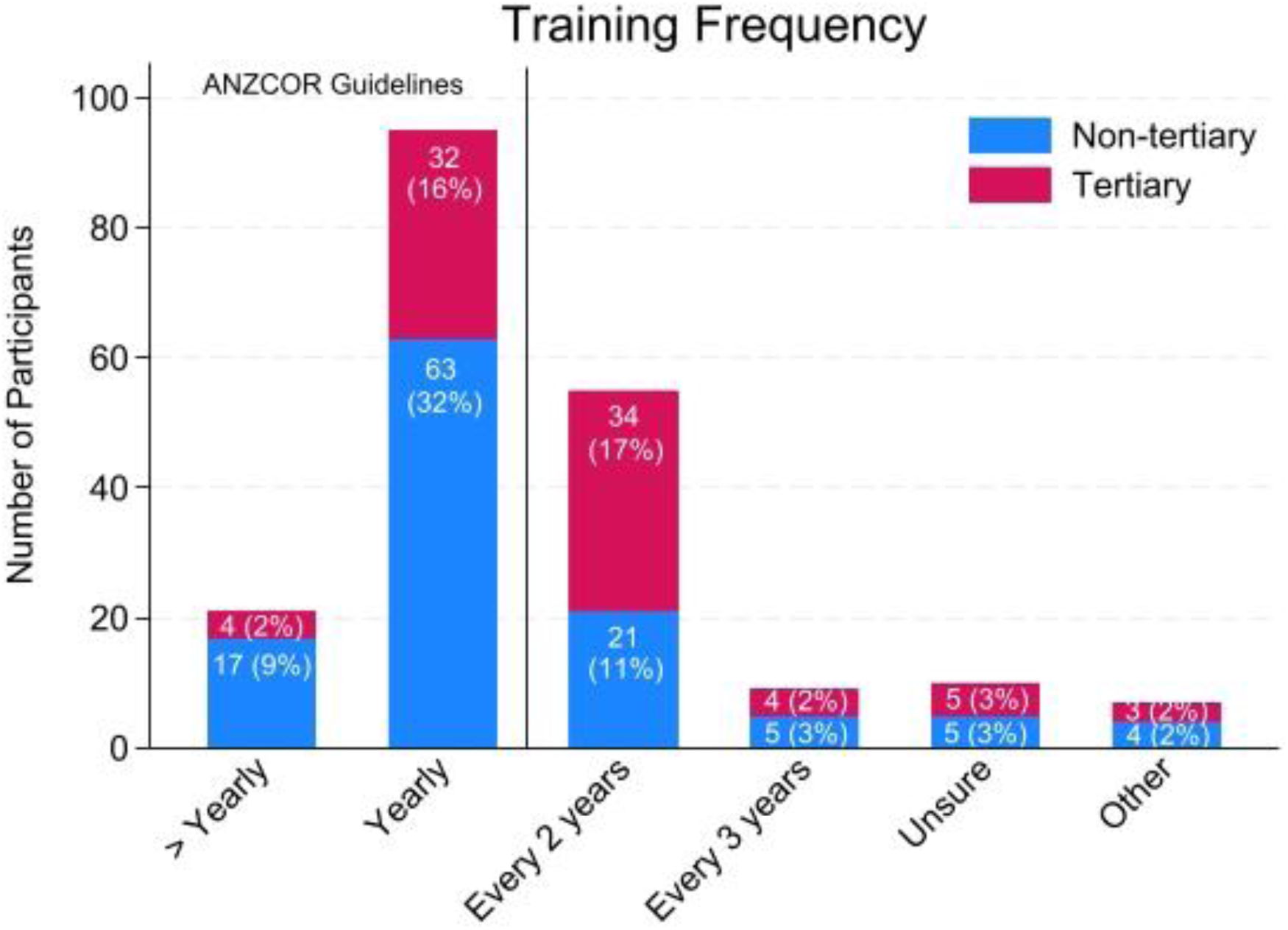
Frequency neonatal resuscitation training is taught in Australian public birthing and newborn care facilities.

**Figure 2:**
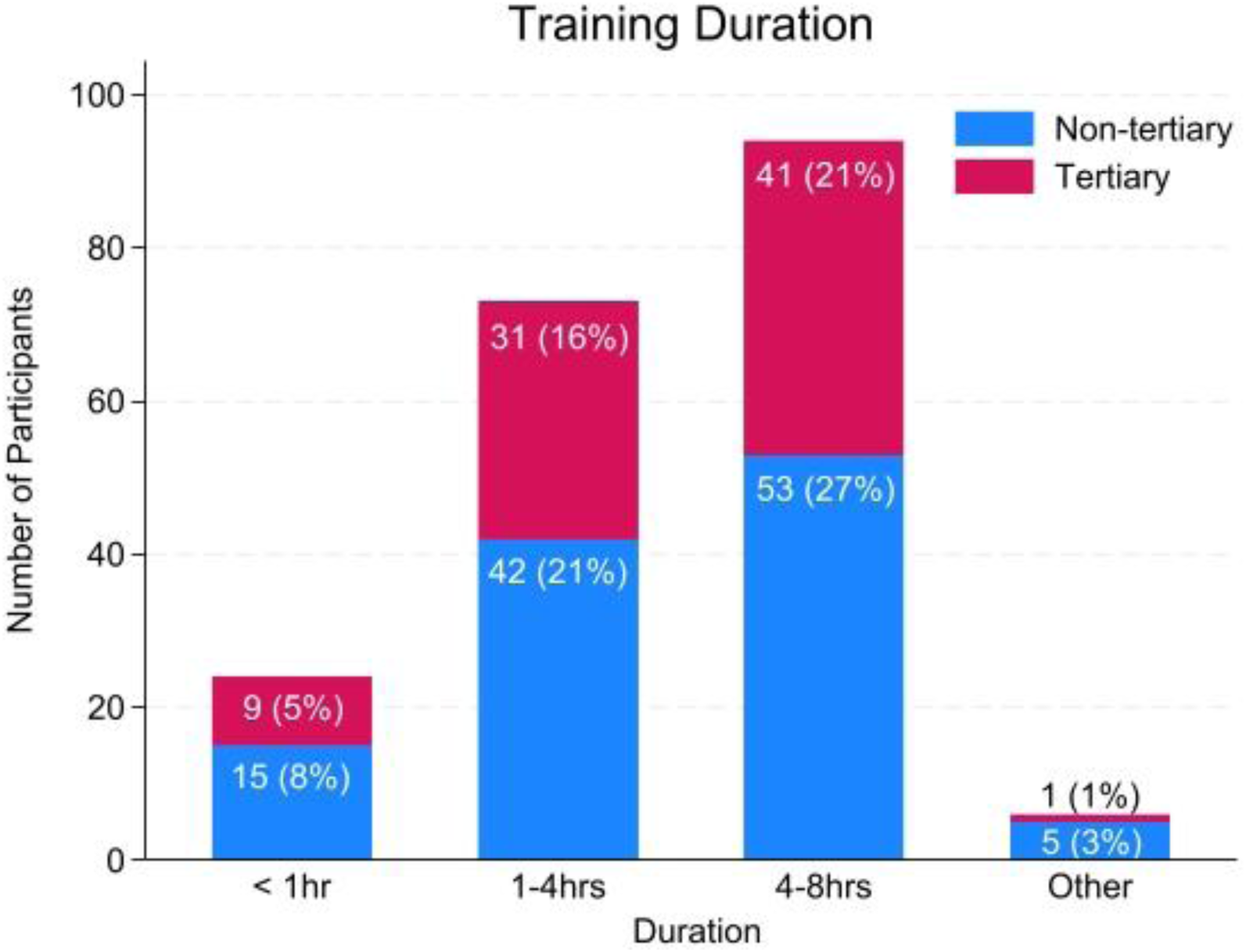
Duration of neonatal resuscitation training taught in Australian public birthing and newborn care facilities.

### Programmes

The training programmes taught in Australian hospitals are presented in Figure 3. All training programs were taught in at least four states or territories with 30% of respondents’ units participating in multiple programmes.

**Figure 3:**
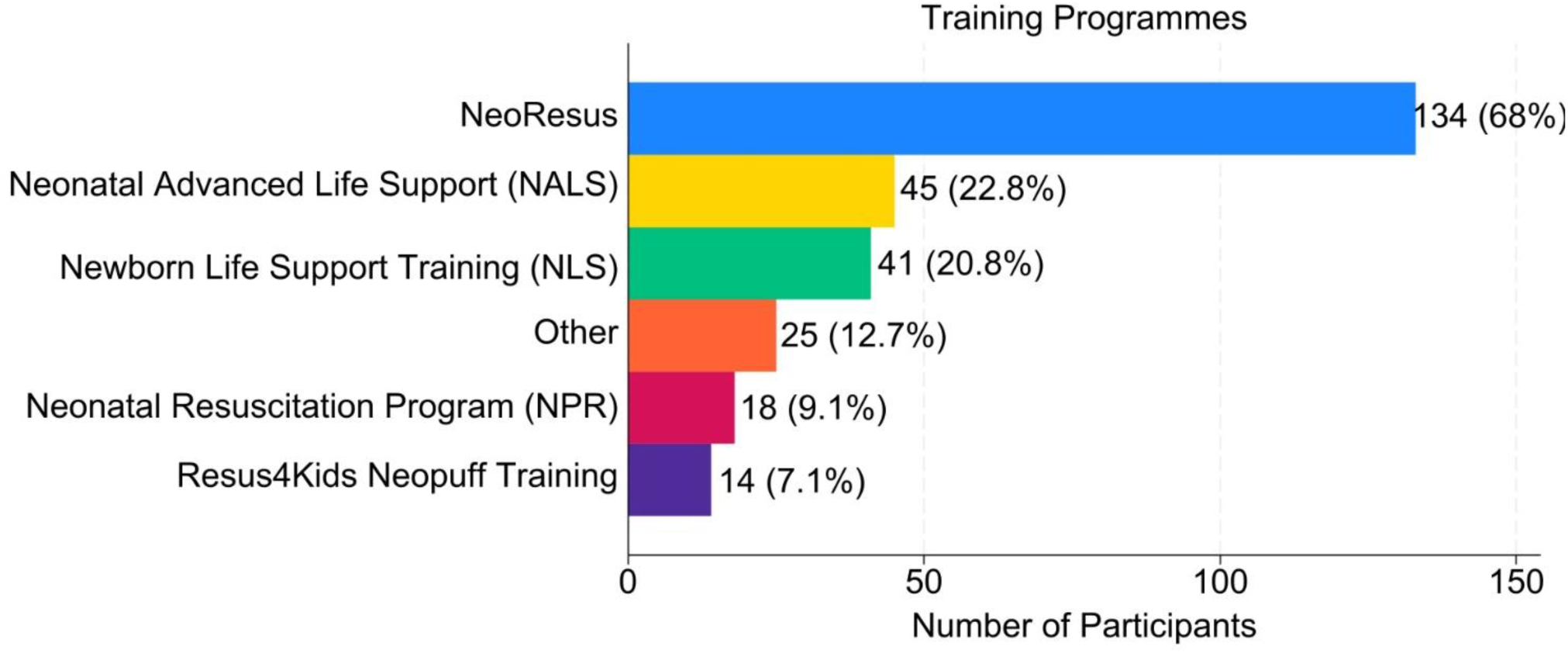
Training programmes taught in Australian public birthing and newborn care facilities. Note: “Other” included internal or regional developed training program.

### Equipment, Simulation and Assessment

Equipment available and used during neonatal resuscitation training is presented in Figure 4. Manikins were available at 195 (99%) respondents neonatal resuscitation training programmes. The most frequent manikin brands used include Laerdal (n=174, 88.3%), Sim characters (n=16, 8.1%) and Ambu (n=11, 5.6%), 68% of them intubatable. Feedback devices used to measure and guide ventilation technique included respiratory function monitors (e.g. ResusRight Juno training monitor) (n=5, 2.5%) or high-tech manikin (e.g. Laerdal QCPR manikin) (n=7, 3.6%).

**Figure 4:**
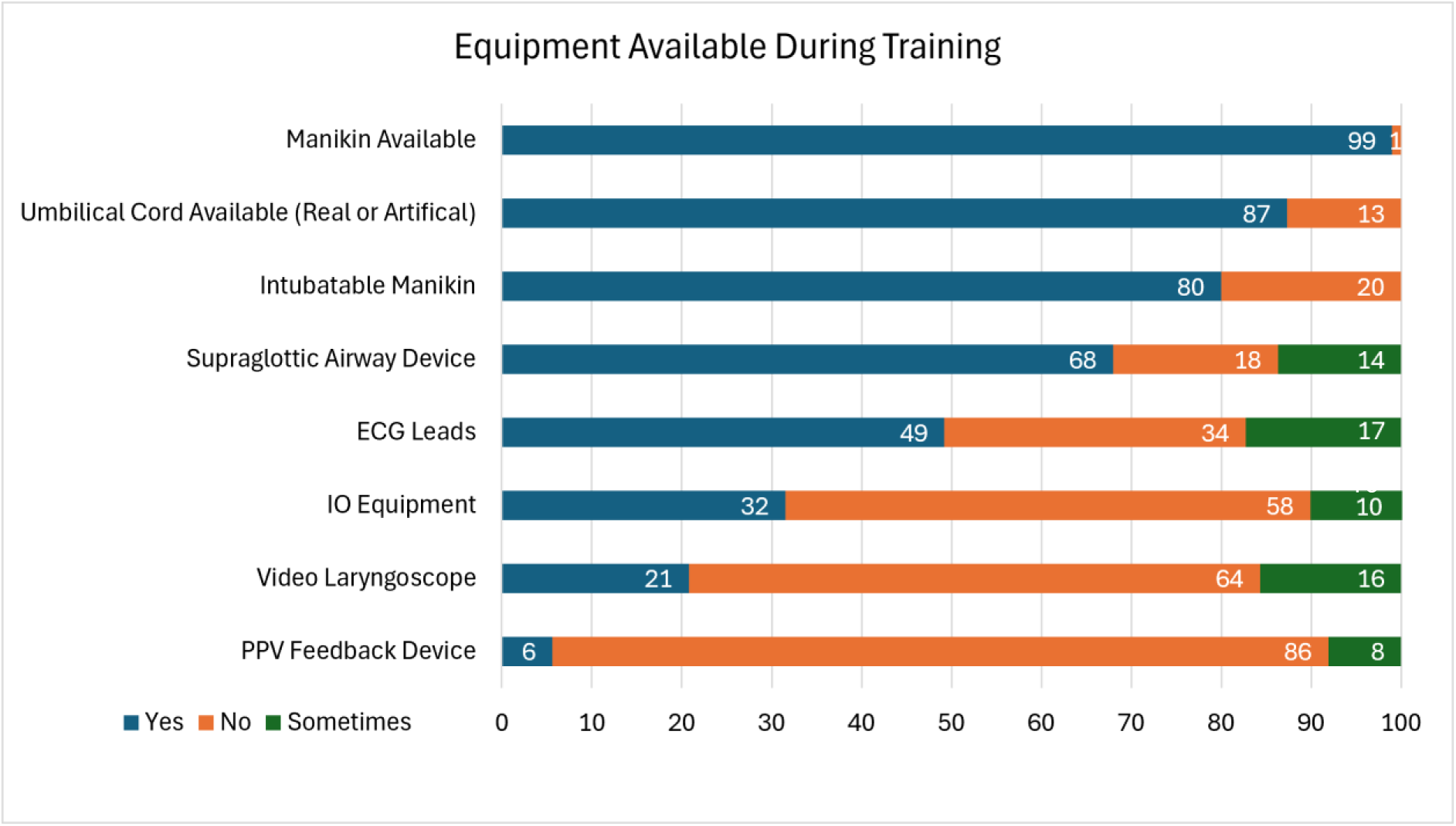
Equipment available and used during neonatal resuscitation training in Australian public birthing and newborn care facilities.

Positive pressure ventilation was taught using self-inflating bags (n=157, 79.7%), t-piece resuscitators (n=196, 99.5%) and flow-inflating bag (n=20, 10.2%). Education in only the TPR was reported by 40 (20.3%) respondents.

Deferred cord clamping was taught regularly in 173 (87.8%) and occasionally in 15 (7.6%) of respondents’ programmes. Cord milking for infants >28weeks was taught regularly in 13 (6.6%) and taught sometimes in 13 (4.1%) of respondents’ programmes.

Simulation based scenarios was used to teach neonatal resuscitation training in 185 (94.4%) respondents programmes. Structured feedback on training programme taught was collected by 161 (81.7%) of respondents’ programmes. The success of training programmes taught were assessed by 185 (93.9%) of respondents’ programmes in the form of a pre-training quiz (n=93, 47.2%), post-training quiz (n=75, 38.1%), certificate of attendance to face-to-face training (n=167, 84.8%) or other (n=27, 13.7%).

### Perceptions of National Standardisation

174 (88.3%) respondents reported a need for the development of a standardised national training program. Of these, 72 (37%) of respondents were from tertiary facilities and 102 (52%) of respondents from non-tertiary facilities.

### Participant Comments

Inductive content analysis of free-text responses identified several themes related to neonatal resuscitation training and national standardisation. Participants frequently described a lack of uniformity across existing training programmes and the absence of formal accreditation pathways, particularly when compared with established adult and paediatric resuscitation programmes. A common concern was the lack of a national instructor credentialing system, which participants perceived contributing to an increased variability in both teaching quality and competency assessment. Limited access to a train the trainer programmes was reported, with participants from rural and remote healthcare facilities describing additional logistic and financial barriers to accessing existing programmes. Strong support was expressed for the development of a national standardised training programme, with cost identified as an important consideration. Participants favoured the development of a government-supported, non-profit delivery model to minimise inequities in access between facilities. Clinicians also advocated for curriculum refinement, including shorter, skills-focused face-to-face training supported by improved online learning, greater emphasis on ventilation and troubleshooting, and scenarios that reflect diverse clinical contexts.

## Discussion

In Australia, most birthing and newborn care facilities have reported providing regular neonatal resuscitation training. Differences were observed in accreditation requirements, training frequency, programme and equipment used to support training delivery. Although many facilities reported practices consistent with national or international recommendations, neonatal resuscitation education is not delivered through a single nationally standardised approach. These findings highlight opportunities to improve consistency in the delivery of neonatal resuscitation education nationally.

In 2005, Foster et al surveyed clinical staff working in birthing facilities across NSW and ACT to examine their experiences with neonatal resuscitation training. One-third of participant from rural and urban non-tertiary units, and 12% of participants in urban tertiary units reported either never having received neonatal resuscitation training or not attended training within the previous two years.^15^ In the present national survey, 12.2% of participants form non-tertiary units and 15.7% of participants from tertiary units reported that neonatal resuscitation training was not conducted at least every two years. Although direct comparison is limited by differences in methodology, the findings suggest improved access to neonatal resuscitation training in non-tertiary settings since the earlier survey. However, a proportion of services continued to report training intervals exceeding two years in the current survey, indicating that practice does not uniformly align with ANZCOR guidelines regarding ongoing education.^7^ This may illustrate the lack of enough human resources to run this programme more frequently.

Variations in guideline recommendations and clinical practice have been reported internationally. A national survey of neonatal resuscitation practices in Spain identified substantial differences between hospitals despite awareness of international resuscitation guidelines.^16^ This suggests that guidelines alone may be insufficient to achieve consistent implementation in practice. An Indian survey of neonatal resuscitation training instructors identified challenges relating to instructor preparation, simulation training and consistency of programme delivery, highlighting the importance of continual instructor development and support in achieving standardised neonatal resuscitation education.^17^

Similar to a 2017 Australian survey of non-tertiary units in NSW and the ACT, participants identified differences in neonatal resuscitation equipment, personnel training and availability of skilled personnel.^18^ Educators recognised instructor preparedness, formal accreditation and equitable access to training programmes for rural and remote services as important considerations for any future national training framework. A strong support for a national standardised neonatal resuscitation training approach was expressed by a large majority of participants (88.3%). This response is consistent with international efforts to improve consistency in neonatal resuscitation education through nationally coordinated training frameworks, both in developed (e.g. NRP and NLS) and developing countries (e.g. Helping Babies Breathe).

Most neonatal resuscitation training programmes taught in Australia have been developed internationally in the United States and United Kingdom for national standardised programmes, with the exception of NeoResus and Resus4Kids which are both Australian developed programmes. The extended use of the programme NeoResus may illustrate a better fit to the Australian levels of care and their diverse birth and newborn care environments. In the current survey, 30% of facilities reported teaching more than one neonatal resuscitation programme. Exposure to multiple programmes may result in differences in educational content leading to disjointed team dynamics, skill quality and conflicting protocols.^19^

Recent guideline updates have introduced recommendations for new technologies and airway management approaches during neonatal resuscitation. ANZCOR most recent guidelines support the use of supraglottic airway as a first response device and an alternative to endotracheal intubation when facemask ventilation is unsuccessful.^7^ Access to supraglottic airways during training was reported by 68.5% of facilities, similar to supraglottic airway use of 67% reported in 2018.^20^ ANZCOR also recommends the use of electrocardiography (ECG) for more rapid and accurate heart rate assessment in the first minutes of life than pulse oximetry alone, where there is availability.^21^ Nearly half of participating facilities (49%) reported training clinicians in the use of ECG during neonatal resuscitation. While survey methodology differs, this represents a notable increase from the limited use of ECG monitoring reported in Australian non-tertiary units in 2017.^18^ More recent ILCOR 2025 recommendations suggest the use of video laryngoscopes for initial intubation attempts where resources permit.^22^ In the current survey, 21-36% of facilities train staff in the use of video laryngoscopy. Collectively, these findings suggest that the implementation of emerging guideline recommendations is occurring, although uptake across Australian services remains variable and would benefit from standardisation. Regular neonatal resuscitation training is important as without ongoing practice, resuscitation knowledge, technical skills and confidence decline over time.^23 24^ Brief refresher activities or re-training at more frequent intervals have been shown to improve skill retention and maintain competency.^25 26^ Inconsistencies in training requirements have previously been identified in Australian maternity services,^9^ suggesting opportunities to strengthen alignment with guideline recommendations through a more coordinated national approach to neonatal resuscitation education.

Blended learning approaches that combine didactic teaching with hands-on simulation have been associated with improved skill acquisition and knowledge retention and are recommended by both ANZCOR and ILCOR.^9 27 25 28^ The use of blended learning approaches aligns with contemporary educational recommendations. Objective feedback technologies, such as respiratory function monitors and high-tech manikins, may enhance acquisition of manual ventilation and psychomotor skills during training.^25 29 30^ Recent ANZCOR and ILCOR guidelines increasingly support the use of feedback technologies during resuscitation training, highlighting opportunities to supplement the predominantly subjective assessment methods reported by survey participants with more objective measures of performance and competency.^22 25^

Although most participating facilities reported using blended learning approaches and simulation-based training, evidence suggests that gaps may persist between training performance and clinical practice.^31 32^ Educational strategies integrating training into the clinical environment, including structured briefings and debriefings, may provide additional opportunities to reinforce both technical and non-technical neonatal resuscitation skills and allow every birth to be a learning event.^25 32^

Variation was observed in the equipment used to support ventilation training. Approximately 20% of facilities reported training staff exclusively with a T-piece resuscitator. While ANZCOR recommends T-piece resuscitators as the preferred device for neonatal positive pressure ventilation, effective use requires access to a compressed gas source and an air-oxygen blender. ^33^ As these resources may not be universally available across all healthcare settings therefore guidelines recommend that a self-inflating bag remains available as a backup device.^18 33^ Training clinicians across a range of ventilation devices is important to ensure preparedness for varying clinical circumstances and equipment availability.

To our knowledge this is the first contemporary national survey describing neonatal resuscitation training practices across Australian public maternal and newborn care facilities. Several limitations should be considered when interpreting these findings. The survey targeted educators at facilities who are involved in neonatal resuscitation training and may therefore underrepresent facilities that do not routinely provide formal training programmes. Findings may be subject to a non-response bias. The survey also did not differentiate between basic and advanced neonatal resuscitation programmes, limiting more detailed comparisons. Finally, variation in local training requirements, including the use of competency assessment rather than formal accreditation may have influenced participant interpretation of some survey questions.

## Conclusion

This study provides the first contemporary national baseline of neonatal resuscitation training across Australian birthing and newborn care facilities. Despite widespread access to training, differences in accreditation requirements, training frequency, programme delivery and educational resources were identified. These findings provide an important evidence base to inform future standardisation efforts and development of a coordinated national approach to neonatal resuscitation education in Australia.

## Data Availability

All data produced in the present study are available upon reasonable request to the authors.

## Contributions

SM survey design, data collection, data analysis, writing of original manuscript draft, manuscript revisions and approval of final manuscript.

MBT survey design, conceptualization, critical review, editing and approval of final manuscript.

MT survey design, conceptualization, critical review, editing and approval of final manuscript.

NW survey design, critical review, editing and approval of final manuscript.

## Acknowledgements

The authors would like to acknowledge Kristina Hall, Krista Lowe and Hannah Skelton for reviewing study survey, providing valuable feedback on its content and design and assistance with compiling the educator contacts list.

We acknowledge funding support from a PhD Research Grant awarded by Cerebral Palsy Alliance (PHD04123) and Australian Government Research Training Program (RTP) Scholarship (N/A) awarded to Stephanie Morakeas. The funders had no role in study design, data collection, data analysis, data interpretation, or writing of the manuscript.

## References

1. Health AIo, Welfare. Australia’s mothers and babies Canberra: AIHW; 2024 [Available from: https://www.aihw.gov.au/reports/mothers-babies/australias-mothers-babies.

2. Boland RA, Davis PG, Dawson JA, et al. Outcomes of infants born at 22–27 weeks’ gestation in Victoria according to outborn/inborn birth status. Arch Dis Child Fetal Neonatal Ed 2017;102(2):F153–F61. doi: 10.1136/fetalneonatal-2015-310313corr1

3. Thompson K, Gardiner J, Resnick S. Outcome of outborn infants at the borderline of viability in Western Australia: a retrospective cohort study. Journal of Paediatrics and Child Health 2016;52(7):728–33. doi: 10.1111/jpc.13187

4. Patel A, Khatib MN, Kurhe K, et al. Impact of neonatal resuscitation trainings on neonatal and perinatal mortality: a systematic review and meta-analysis. BMJ Paediatr Open 2017;1(1) doi: 10.1136/bmjpo-2017-000183

5. Dempsey E, Joyce R, Neveln N, et al. Standardised formal resuscitation training programmes for reducing mortality and morbidity in newborn infants. Cochrane Database Syst Rev 2026;4(4):Cd009106. doi: 10.1002/14651858.CD009106.pub3 [published Online First: 20260402]

6. Perlman JM, Wyllie J, Kattwinkel J, et al. Part 7: Neonatal Resuscitation: 2015 International Consensus on Cardiopulmonary Resuscitation and Emergency Cardiovascular Care Science With Treatment Recommendations. Circulation 2015;132(16 suppl 1):S204–S41. doi: 10.1161/cir.0000000000000276

7. (ANZCOR) AaNZCoR. ANZCOR Guideline 13.5 – Tracheal Intubation and Ventilation of the Newborn, 2021.

8. Guidlines QC. Queensland Clinical Guidlines - Neonatal Resuscitation 2022 [Available from: https://www.health.qld.gov.au/data/assets/pdf_file/0011/140600/g-resus.pdf.

9. Williams J, Ebert L, Duff J. Neonatal resuscitation training for midwives in Australia: A discussion of current practice. Women and Birth 2020;33(6):e505–e10. doi: 10.1016/j.wombi.2020.01.002

10. RANZCORG TRANZCoOaG-. Training sites and units - Accreditation 2024 [Available from: https://ranzcog.edu.au/training-topics/training-sites/ accessed 23.12.2024 2024.

11. Foster K, Craven P, Reid S. Neonatal resuscitation educational experience of staff in New South Wales and Australian Capital Territory hospitals. Journal of paediatrics and child health 2006;42(1-2):16–19. doi: 10.1111/j.1440-1754.2006.00783.xDigital

12. Sharma A, Minh Duc NT, Luu Lam Thang T, et al. A Consensus-Based Checklist for Reporting of Survey Studies (CROSS). J Gen Intern Med 2021;36(10):3179–87. doi: 10.1007/s11606-021-06737-1 [published Online First: 20210422]

13. Health NMo. NSW Health Guide to Role Delineation of Clinical Services. In: Health N, ed., 2024.

14. Homer CS, Biggs J, Vaughan G, et al. Mapping maternity services in Australia: location, classification and services. Aust Health Rev 2011;35(2):222–29. doi: 10.1071/AH10908

15. Foster K, Craven P, Reid S. Neonatal resuscitation educational experience of staff in New South Wales and Australian Capital Territory hospitals. J Paediatr Child Health 2006;42(1-2):16–9. doi: 10.1111/j.1440-1754.2006.00783.x

16. Iriondo M, Thió M, Burón E, et al. A survey of neonatal resuscitation in Spain: gaps between guidelines and practice. Acta Paediatr 2009;98(5):786–91. doi: 10.1111/j.1651-2227.2009.01233.xDigital

17. Patel P, Nimbalkar S, Shinde M. Insights from a cross-sectional survey of neonatal resuscitation instructors from India. Sci Rep 2023;13(1):15255. doi: 10.1038/s41598-023-42382-w

18. Shah D, Kalpesh J, Tracy M, et al. Survey of neonatal resuscitation equipment and training in NSW/ACT non-tertiary birthing units. Journal of Paediatrics and Child Health 2017;53(52) doi: http://dio.org/10.1111/jpc.13494_262

19. Schierholz E, Wetzel EA, Thomas AR, et al. Resuscitation education for NICU providers: Current practice and recommendations for NRP and PALS in the NICU. Semin Perinatol 2024;48(8):151991. doi: 10.1016/j.semperi.2024.151991 [published Online First: 20241009]

20. Goel D, Shah D, Hinder M, et al. Laryngeal mask airway use during neonatal resuscitation: a survey of practice across newborn intensive care units and neonatal retrieval services in Australian New Zealand Neonatal Network. J Paediatr Child Health 2020;56(9):1346–50. doi: http://dio.org/10.1111/jpc.15110

21. (ANZCOR) AaNZCoR. ANZCOR Guideline 13.3 – Assessment of the Newborn. ANZCOR.org: ANZCOR, 2021.

22. Berg KM, Bray JE, Djärv T, et al. Executive Summary: 2025 International Liaison Committee on Resuscitation Consensus on Science With Treatment Recommendations. Resuscitation 2025;215 doi: 10.1016/j.resuscitation.2025.110805

23. Wyllie J, Perlman JM, Kattwinkel J, et al. Part 7: Neonatal resuscitation: 2015 International Consensus on Cardiopulmonary Resuscitation and Emergency Cardiovascular Care Science with Treatment Recommendations. Resuscitation 2015;95:e169–201. doi: 10.1016/j.resuscitation.2015.07.045

24. Caldelari M, Floris L, Marchand C, et al. Maintaining the knowledge and neonatal resuscitation skills of student midwives 6 months after an educational program. Arch Pediatr 2019;26(6):385–92. doi: 10.1016/j.arcped.2019.05.001

25. ANZCOR. Guideline 10.2 – Advanced Life Support (ALS) Training 2026 [Available from: https://www.anzcor.org/home/education-and-implementation/guideline-10-2-advanced-life-support-als-training.

26. Haynes JC, Rettedal SI, Ushakova A, et al. How Much Training Is Enough? Low-Dose, High-Frequency Simulation Training and Maintenance of Competence in Neonatal Resuscitation. Simul Healthc 2024;19(6):341–49. doi: http://dio.org/10.1097/sih.0000000000000783 [published Online First: 20240306]

27. Rheinberger J, Curtis K, McCloughen A, et al. What are the training needs of emergency department resuscitation nurses? A scoping review. Australasian Emergency Care 2024;27(4):244–53. doi: 10.1016/j.auec.2024.06.001

28. Wyckoff MH, Greif R, Morley PT, et al. 2022 International Consensus on Cardiopulmonary Resuscitation and Emergency Cardiovascular Care Science With Treatment Recommendations: Summary From the Basic Life Support; Advanced Life Support; Pediatric Life Support; Neonatal Life Support; Education, Implementation, and Teams; and First Aid Task Forces. Circulation 2022;146(25):e483–e557. doi: 10.1161/CIR.0000000000001095

29. O’Currain E, Thio M, Dawson JA, et al. Respiratory monitors to teach newborn facemask ventilation: a randomised trial. Arch Dis Child Fetal Neonatal Ed 2019 doi: http://dio.org/10.1136/archdischild-2018-316118 [published Online First: 2019/01/15]

30. Ades A, Lee HC. Update on simulation for the Neonatal Resuscitation Program. Semin Perinatol 2016;40(7):447–54. doi: 10.1053/j.semperi.2016.08.005

31. Raghunathan K, East C, Poudel K. Barriers and enablers for implementation of clinical practice guidelines in maternity and neonatal settings: A rapid review. PLoS One 2024;19(12):e0315588. doi: 10.1371/journal.pone.0315588 [published Online First: 20241216]

32. Bettinger K, Mafuta E, Mackay A, et al. Improving Newborn Resuscitation by Making Every Birth a Learning Event. Children 2021;8(12):1194. doi: 10.3390/children8121194

33. (ANZCOR) AaNZCoR. ANZCOR Guideline 13.4 – Airway Management and Mask Ventilation of the Newborn. ANZCOR.org: ANZCOR, 2021.

