## Supplementary material for "Neonatal Resuscitation Training in Australian Public Birthing and Newborn Care Facilities: A National Cross-Sectional Survey": Supplemmentary

#### Questionnaire:

*Confidential*



### National Newborn Resuscitation Training Survey for Educators

Dear Colleagues,

The PSANZ Newborn Resuscitation and Stabilisation Subcommittee (NR&S) recognises the critical role that effective newborn resuscitation practices play in improving neonatal outcomes.

Evidence from other jurisdictions demonstrates that standardised training programs significantly enhance the quality of resuscitation efforts, reduce variability in practice, and lead to better health outcomes for newborns. This survey represents a valuable step toward achieving similar advancements in our region.

In this survey we aim to gather knowledge on key aspects of newborn resuscitation, including training requirements, staffing, and equipment used in training, with the ultimate goal of informing the development of a standardised newborn resuscitation training program in Australia.

Answering this anonymous survey will take up to 10 minutes.

Please note all questions are in regard to newborn resuscitation training only.

By participating you consent to the use of your response data for the study.

Thank you!

##### Section 1: Background

1. What state in Australia are is your workplace?
   - ◯ NSW
   - ◯ ACT
   - ◯ VIC
   - ◯ SA
   - ◯ WA
   - ◯ NT
   - ◯ QLD
   - ◯ TAS

2. What type of neonatal site is your workplace?
   - ◯ Level 1 (No planned maternity/neonatal service (NPS))
   - ◯ Level 2 (newborns >=37 weeks)
   - ◯ Level 3 (newborns >=34 weeks)
   - ◯ Level 4 (newborns >=32 weeks)
   - ◯ Level 5 (intensive care for critically ill newborns no extreme preterm infants)
   - ◯ Level 6 (neonatal surgical and medical subspecialty services)

3. What is your role?
   - ◯ Senior medical staff (consultant level)
   - ◯ Junior medical staff (residents, registrars and fellows)
   - ◯ Nursing staff
   - ◯ Midwifery staff
   - ◯ Other

3a. Please specify "other"

__________________________________

3b. Please specify senior medical staff role:
   - ◯ Neonatologist
   - ◯ Paediatrician
   - ◯ Obstetrician
   - ◯ General Practitioner
   - ◯ Anaesthetist
   - ◯ Other

| | | |
|---|---|---|
| 3c. | Please specify junior medical staff role: | ◯ Paediatric trainee<br>◯ Obstetric trainee<br>◯ Other |
| 3d. | Please specify nursing roles: | ◯ Paediatric<br>◯ General<br>◯ Nurse practitioner<br>◯ Nurse Educator<br>◯ Other |
| 4. | Do you job share this role with someone else? | ◯ Yes<br>◯ No<br>◯ Other |
| 4a. | Please specify "other" | __________________________________ |
| 4b. | If "yes", who do you share this role with? | ◯ Nursing<br>◯ Medical<br>◯ Other |

**Section 2: Education and Training**

| | | |
|---|---|---|
| 5. | Is an accreditation in neonatal resuscitation required at your site? | ◯ Yes<br>◯ No<br>◯ Unsure |
| 6. | What is the required training interval required by your institution? | ◯ < Once a year<br>◯ Yearly<br>◯ Every 2 years<br>◯ Every 3 years<br>◯ Unsure<br>◯ Other |
| 6a. | Please specify "other" | __________________________________ |
| 7. | Where do staff attend training?<br>Select all that apply | ☐ Onsite<br>☐ Offsite<br>☐ Online Only<br>☐ Unsure |
| 8. | Who is responsible for facilitating the training program? | ◯ Doctors / Nurse Practitioners<br>◯ Nurses / Midwives<br>◯ Both<br>◯ Other |
| 8a. | Please specify "other" | __________________________________ |
| 9. | Is the training program attended by? | ◯ Doctors / Nurse Practitioners<br>◯ Nurses / Midwives<br>◯ Both<br>◯ Other |
| 9a. | Please specify "other" | |

| | | |
|---|---|---|
| 10. | Is the educator: | ◯ Internal<br>◯ External<br>◯ Both<br>◯ Unsure |
| 11. | What type of training program is taught at your institution?<br>Select all that apply | ☐ NeoResus<br>☐ NRP<br>☐ NLS<br>☐ NALS<br>☐ Resus4Kids neopuff training<br>☐ Other |
| 11a. | Please specify "other" | __________________________________ |
| 12. | What is the typical length of face- to- face training session?<br>Select all that apply | ☐ < 1hr<br>☐ 1-4hr<br>☐ 4-8hr<br>☐ Other |
| 12a. | Please specify "Other" | __________________________________ |
| 13. | Does the training course require reading or course work prior to attendance? | ◯ Yes<br>◯ No<br>◯ Unsure |

**Recent ILCOR recommendations for newborn resuscitation training suggest improved training practices. Please answer the following questions based on your current training practices:**

| | | |
|---|---|---|
| 14. | Is training performed using a:<br><br>Select all that apply | ☐ Self-Inflating Bag<br>☐ T-Piece Resuscitator (eg. NeoPuff)<br>☐ Flow-inflating bag/Anesthesia bag<br>☐ Other |
| 14a. | Please specify "Other" | __________________________________ |
| 15. | What brand/s of manikins do you use during resuscitation training?<br>Select all that apply | ☐ Laerdal<br>☐ TrueCorp<br>☐ Gaumard Scientific<br>☐ Somso<br>☐ Nasco Life/Form<br>☐ Ambu<br>☐ Lifecast<br>☐ Sim characters<br>☐ Medical X<br>☐ Syndaver Labs<br>☐ Medvision<br>☐ CAE<br>☐ Other |
| 15a. | Please specify "Other" | __________________________________ |

16. Are intubatable or non-intubatable manikins used during resuscitation training?
   - ◯ Intubatable
   - ◯ Non-intubatable
   - ◯ Both
   - ◯ Unsure

17. Are ECG leads use during newborn resuscitation discussed in training?
   - ◯ Yes
   - ◯ No
   - ◯ Sometimes

17a. Comment: __________________________________

18. Is training performed for supraglottic airway insertion?
   - ◯ Yes
   - ◯ No
   - ◯ Sometimes

18a. Comment: __________________________________

18b. If 'yes', please list brand __________________________________

19. Is a video laryngoscope used during training?
   - ◯ Yes
   - ◯ No
   - ◯ Sometimes

19a. Comment: __________________________________

20. What type of umbilical cords are used during training?
   - ◯ Real
   - ◯ Artificial
   - ◯ Both
   - ◯ Unsure

20a. Comment: __________________________________

21. Is delayed cord clamping taught?
   - ◯ Yes
   - ◯ No
   - ◯ Sometimes

21a. Comment: __________________________________

22. Is cord milking taught for infants >28 weeks?
   - ◯ Yes
   - ◯ No
   - ◯ Sometimes

22a. Comment: __________________________________

23. Is training performed on Intraosseous (IO) insertion?
   - ◯ Yes
   - ◯ No
   - ◯ Sometimes

24a. Comment: __________________________________

25. Are respiratory function monitors (RFMs) used during resuscitation training to provide measured pressure or tidal volume feedback (eg. ResusRight/Monivent/Laerdal QCPR Feedback)?
- ◯ Yes
- ◯ No
- ◯ Sometimes

25a. Comment: __________________________________

25b. If 'yes', please specify brand (eg.ResusRight/Monivent/Laerdal QCPR Feedback)? __________________________________

26. Do you practice simulation based scenarios in your newborn resuscitation?
- ◯ Yes
- ◯ No
- ◯ Sometimes

26a. Comment: __________________________________

#### Section 3: Evaluation of Current Training Programs

27. Do you collect structured feedback on your training program?
- ◯ Yes
- ◯ No
- ◯ Sometimes

29. Do you see a need or benefits for a standardised national training program?
- ◯ Yes
- ◯ No
- ◯ Unsure

28. Do you have a way to assess success of resuscitation training?
Select all that apply
- ☐ Manikin/equipment feedback (eg. QCPR manikin, RFM or other)
- ☐ Pre-training quiz
- ☐ Post-training quiz
- ☐ Certificate of attendance to face-to-face training
- ☐ Other

28a. Please specify "other" __________________________________

29. We welcome any other comments: __________________________________

**Appendix Table 1:**

| **Training Accreditation Requirement** | **Number (%)** |
|---|---|
| Yes | 173 (87.8%) |
| No | 12 (6.1%) |
| Unsure | 12 (6.1%) |
| **Training Location*** | |
| Onsite | 190 (96.4%) |
| Offsite | 26 (13.2%) |
| Online | 54 (27.4%) |
| Unsure | 1 (0.5%) |
| **Training Facilitators** | |
| Doctors/Nurse Practitioners | 10 (5.1%) |
| Nurses/Midwives | 64 (32.5%) |
| Both | 100 (50.8%) |
| Other | 23 (11.7%) |
| **Facilitator Origin** | |
| Internal | 192 (97.5%) |
| External | 29 (14.7%) |
| **Training Attendees** | |
| Doctors/Nurse Practitioners | 0 (0%) |
| Nurses/Midwives | 21 (10.7%) |
| Both | 155 (78.7%) |
| Other | 21 (10.7%) |
| **Length of face-to-face Training** | |
| < 1 hr | 24 (12.2%) |
| 1-4 hrs | 73 (37.1%) |
| 4-8 hrs | 94 (47.7%) |
| Other | 6 (3.0%) |
| **Pre-course work required** | |
| Yes | 181 (91.9%) |
| No | 15 (7.6%) |
| Unsure | 1 (0.5%) |
| **Training Frequency** | |
| More than once yearly | 21 (10.7%) |
| Yearly | 95 (48.2%) |
| Every 2 years | 55 (27.9%) |
| Every 3 years | 9 (4.6%) |
| Unsure | 10 (5.1%) |
| Other | 7 (3.6%) |
| *Multiple answers were allowed for this survey question. | |

**Appendix Table 2:**

| Manikin Brands | Number (%) |
|---|---|
| Laerdal | 174 (88.3%) |
| TrueCorp | 1 (0.5%) |
| Gaumard Scientific | 4 (2.0%) |
| Somso | 1 (0.5%) |
| Nasco Life/Form | 4 (2.0%) |
| Ambu | 11 (5.6%) |
| Lifecast | 5 (2.5%) |
| Sim Characters | 16 (8.1%) |
| Medical X | 0 (0%) |
| Syndaver Labs | 0 (0%) |
| Medvision | 1 (0.5%) |
| CAE | 0 (0%) |
| No Manikin Available | 2 (1.0%) |
| Other | 24 (12.2%) |

**Appendix Table 3: Participants Demographics, Maternity/Neonatal Service Level by State.**

| State | Maternity/Neonatal Level | Number (%) |
|---|---|---|
| QLD | Level 1 (No planned maternity/neonatal service) | 1 (0.5%) |
| | Level 2 (newborns >=37 weeks) | 12 (6.1%) |
| | Level 3 (newborns >=34 weeks) | 1 (0.5%) |
| | Level 4 (newborns >=32 weeks) | 21 (10.7%) |
| | Level 5 (intensive care for critically ill newborns including extreme preterm infants) | 7 (3.6%) |
| | Level 6 (neonatal surgical and medical subspeciality services) | 14 (7.1%) |
| | Total | 56 (28.4%) |
| NSW | Level 1 (No planned maternity/neonatal service) | 1 (0.5%) |
| | Level 2 (newborns >=37 weeks) | 10 (5.1%) |
| | Level 3 (newborns >=34 weeks) | 12 (6.1%) |
| | Level 4 (newborns >=32 weeks) | 7 (3.6%) |
| | Level 5 (intensive care for critically ill newborns including extreme preterm infants) | 12 (6.1%) |
| | Level 6 (neonatal surgical and medical subspeciality services) | 12 (6.1%) |
| | Total | 54 (27.4%) |
| VIC | Level 1 (No planned maternity/neonatal service) | 4 (2.0%) |
| | Level 2 (newborns >=37 weeks) | 4 (2.0%) |
| | Level 3 (newborns >=34 weeks) | 6 (3.0%) |
| | Level 4 (newborns >=32 weeks) | 17 (8.6%) |
| | Level 5 (intensive care for critically ill newborns including extreme preterm infants) | 5 (2.5%) |
| | Level 6 (neonatal surgical and medical subspeciality services) | 5 (2.5%) |
| | Total | 41 (20.8%) |
| WA | Level 1 (No planned maternity/neonatal service) | 0 (0%) |
| | Level 2 (newborns >=37 weeks) | 4 (2.0%) |
| | Level 3 (newborns >=34 weeks) | 4 (2.0%) |
| | Level 4 (newborns >=32 weeks) | 1 (0.5%) |

| | | |
|---|---|---|
| | Level 5 (intensive care for critically ill newborns including extreme preterm infants) | 6 (3.0%) |
| | Level 6 (neonatal surgical and medical subspeciality services) | 4 (2.0%) |
| | Total | 19 (9.6%) |
| SA | Level 1 (No planned maternity/neonatal service) | 0 (0.5%) |
| | Level 2 (newborns >=37 weeks) | 2 (1.0%) |
| | Level 3 (newborns >=34 weeks) | 4 (2.0%) |
| | Level 4 (newborns >=32 weeks) | 1 (0.5%) |
| | Level 5 (intensive care for critically ill newborns including extreme preterm infants) | 2 (1.0%) |
| | Level 6 (neonatal surgical and medical subspeciality services) | 6 (3.0%) |
| | Total | 15 (7.6%) |
| TAS | Level 1 (No planned maternity/neonatal service) | 0 (0%) |
| | Level 2 (newborns >=37 weeks) | 0 (0%) |
| | Level 3 (newborns >=34 weeks) | 1 (0.5%) |
| | Level 4 (newborns >=32 weeks) | 2 (1.0%) |
| | Level 5 (intensive care for critically ill newborns including extreme preterm infants) | 2 (1.0%) |
| | Level 6 (neonatal surgical and medical subspeciality services) | 0 (0%) |
| | Total | 5 (2.5%) |
| NT | Level 1 (No planned maternity/neonatal service) | 0 (0%) |
| | Level 2 (newborns >=37 weeks) | 0 (0%) |
| | Level 3 (newborns >=34 weeks) | 0 (0%) |
| | Level 4 (newborns >=32 weeks) | 0 (0%) |
| | Level 5 (intensive care for critically ill newborns including extreme preterm infants) | 2 (1.0%) |
| | Level 6 (neonatal surgical and medical subspeciality services) | 2 (1.0%) |
| | Total | 4 (2.0%) |
| ACT | Level 1 (No planned maternity/neonatal service) | 0 (0%) |
| | Level 2 (newborns >=37 weeks) | 0 (0%) |
| | Level 3 (newborns >=34 weeks) | 0 (0%) |
| | Level 4 (newborns >=32 weeks) | 0 (0%) |
| | Level 5 (intensive care for critically ill newborns including extreme preterm infants) | 3 (1.5%) |
| | Level 6 (neonatal surgical and medical subspeciality services) | 0 (0%) |
| | Total | 3 (1.5%) |
| | Total | 197 |
